# Impact of COVID-19 and Vaccination on Lower Urinary Tract Symptoms: Insights from a Prospective Cohort Study

**DOI:** 10.1101/2025.05.12.25327476

**Authors:** Julia Souza, Jose de Bessa, Natássia Truzzi, Carolina Rocha, Bruno Araujo, Julyana Moromizato, Thulio Brandão, Rachel Mazoni, Marcelo Hisano, Zein M. Sammour, Homero Bruschini, William C. Nahas, Cristiano M. Gomes

**Affiliations:** Division of Urology, University of Sao Paulo School of Medicine, Sao Paulo, Brazil; Department of Surgery, State University of Feira de Santana, Bahia, Brazil

**Keywords:** Lower urinary tract symptoms, COVID-19, Overactive bladder, Urinary incontinence, Vaccination, SARS-CoV-2

## Abstract

**Aims:** To evaluate the prevalence, clinical course, and risk factors of lower urinary tract symptoms (LUTS) in patients hospitalized with COVID-19, and to assess associations with comorbidities, disease severity, and vaccination status.

**Methods:** We conducted a prospective cohort study of adult, non-ICU patients hospitalized with confirmed COVID-19 between July 2021 and March 2022. LUTS were assessed using the IPSS, ICIQ-OAB, and ICIQ-UI SF questionnaires during hospitalization, and at one and three months post-discharge. Associations with sex, comorbidities, COVID-19 severity, and vaccination status were analyzed using multivariable logistic regression.

**Results:** Among 168 patients (55.4% male, median age 58 years), 31.0% had moderate to severe LUTS during hospitalization, with storage symptoms predominating. Overactive bladder symptoms were present in 36.7%, and urinary incontinence in 34.5%, more frequently among women. At three months, moderate to severe LUTS declined to 21.9%, and both OAB and UI also decreased significantly. No associations were found between LUTS and comorbidities or disease severity. Fully vaccinated patients had higher odds of moderate to severe LUTS during hospitalization (adjusted OR 10.56, 95% CI 4.13–26.9), particularly those vaccinated with inactivated virus vaccines (BBIBP-CorV).

**Conclusions:** LUTS are prevalent in the acute phase of COVID-19, especially among women, but tend to improve within three months. Unexpectedly, full vaccination— especially with inactivated virus vaccines—was associated with increased odds of moderate to severe LUTS during hospitalization. Further studies are warranted to explore the underlying mechanisms and long-term implications.

## INTRODUCTION

The COVID-19 pandemic has had a profound impact on global health, leading to widespread hospitalizations and significant mortality. While primarily characterized by respiratory symptoms, the disease caused by SARS-CoV-2 has demonstrated a capacity to affect multiple organ systems, including the gastrointestinal tract, liver, bone marrow, kidneys, and bladder [1]. It has been shown that COVID-19 can also cause de novo lower urinary tract symptoms (LUTS)—such as increased urinary frequency, urgency, and nocturia—or exacerbate pre-existing LUTS [2–7]. These symptoms are thought to arise through mechanisms that may include viral cystitis, systemic inflammation, or alterations in autonomic function [7].

Early observations noted urinary frequency as a potential symptom of COVID-19, prompting recommendations to include it in initial clinical evaluations. A Turkish study of 96 hospitalized COVID-19 patients demonstrated an increase in storage LUTS in both sexes, with women also reporting high rates of stress urinary incontinence during hospitalization [2]. Similarly, a survey of post-hospitalization patients identified new-onset LUTS in 39 individuals, with urinary frequency and nocturia reported by 85% and 87%, respectively [3]. Despite these findings, the overall prevalence of LUTS during the acute phase of COVID-19 remains poorly understood.

Beyond the acute phase, a high prevalence of LUTS has been observed among long COVID patients. A six-month follow-up study of post-hospitalization individuals revealed that 42.4% experienced moderate to severe LUTS, with nearly a quarter reporting a significant impact on their quality of life (QoL) [8]. However, the trajectory of LUTS from the acute phase to recovery is unclear, as is the role of vaccination and emerging SARS-CoV-2 variants in modifying these symptoms.

While the critical phase of the pandemic has subsided, COVID-19 persists globally and remains a potential threat, particularly with the emergence of new variants and the possibility of future surges [9, 10]. Understanding its broader health impact is essential - especially in relation to LUTS, which can significantly impair QoL. A recent expert panel emphasized the need for studies evaluating COVID-19’s effects on the urinary tract, including the relationship between disease severity, vaccination, and the progression of LUTS [11].

This study aims to investigate the prevalence of LUTS during the acute phase of COVID-19 in a hospitalized population and track their progression in the short to mid-term post-discharge. Additionally, we explore the impact of pre-existing comorbidities, disease severity, and vaccination status on LUTS development.

## MATERIALS AND METHODS

### Study Design and Setting

This prospective cohort study was conducted at a university-affiliated tertiary hospital. Patients were enrolled between July 2021 and March 2022.

### Participants

Adult men and women (≥18 years) hospitalized with laboratory-confirmed COVID-19, who were not in intensive care were included. Eligibility required willingness to participate and provision of written informed consent. Kidney transplant recipients were included if transplanted at least six months earlier. Patients with urinary tract infections or other urological conditions that could confound the assessment of LUTS were excluded. These conditions included oligoanuria, bladder or prostate cancer and neurological diseases.

### Preexisting comorbidities, parameters of COVID-19 severity and vaccination status

Clinical data were collected, including functional status at the time of the initial assessment (need for supplemental oxygen, mobility status, and method of urination—spontaneous or via bladder catheterization) and pre-existing comorbidities such as diabetes mellitus, systemic arterial hypertension (SAH), obesity (body mass index >30), and prior kidney transplantation. Additionally, key hospitalization details, including the requirement for intensive care or orotracheal intubation before enrollment and the extent of lung damage observed on chest computed tomography (CT), were documented as indicators of COVID-19 severity.

Patients were classified as fully vaccinated if they had received both doses of a two-dose vaccine or a single dose of a one-dose vaccine.

### Prevalence of LUTS and impact on quality of life

During hospitalization for acute COVID-19, participants completed validated versions of the International Prostatic Symptom Score (IPSS) [12], the International Consultation on Incontinence Questionnaire Overactive Bladder (ICIQ-OAB) [13], and the ICIQ – Urinary Incontinence Short Form (ICIQ-UI SF) [14] to assess the prevalence of LUTS, overactive bladder (OAB), and urinary incontinence (UI).

The IPSS assessed specific symptoms, including frequency, nocturia, incomplete emptying, intermittency, slow stream, and straining. Symptoms were considered present if they occurred less than half the time or more. The severity of LUTS was categorized as mild (IPSS ≤7), moderate (IPSS 8–19), or severe (IPSS >19).

The ICIQ-OAB was used to evaluate OAB symptoms, with severity rated on a scale of 0–4. Patients with a score ≥1 on the incontinence question were classified as having urgency urinary incontinence (UUI). Individuals with a total score ≥3 who reported urgency and/or UUI were identified as having OAB.

The ICIQ-UI SF evaluated urinary incontinence, its severity, and associated bother, including stress urinary incontinence (SUI) (leakage during coughing, sneezing, or physical activity), UUI (leaking before reaching the toilet) and post-micturition dribbling (leakage after urination while dressing).

Multiple questionnaires assessed urgency and UUI; the results reported were obtained using the ICIQ-OAB, prioritizing its specific design for these symptoms.

To differentiate preexisting LUTS from de novo symptoms, patients were asked whether they had experienced LUTS prior to contracting COVID-19. Those reporting previous symptoms were questioned about whether their symptoms had worsened, improved, or remained unchanged.

Quality of life related to LUTS was evaluated using the IPSS QoL question, with impairment defined as a score ≥4 (―mostly dissatisfied‖ or worse).

We assessed whether preexisting comorbidities, COVID-19 severity indicators, and vaccination status were associated with the presence and severity of LUTS. **Follow-Up Assessments:** The research team contacted participants one month and three months after discharge to monitor changes in LUTS over time. At each follow-up, patients completed the same validated questionnaires administered at baseline.

### Ethical Considerations

The study was approved by the local ethics committee (approval number 4.765.252). Informed consent was obtained from all participants prior to their inclusion in the study.

## Statistical Analysis

Quantitative variables were expressed as medians and interquartile ranges (IQRs), while qualitative variables were expressed as absolute values, percentages, or proportions. Continuous variables were compared using Student’s t-test or ANOVA, depending on the number of groups. Categorical variables were compared using the Chi-squared test or Fisher’s exact test when appropriate. Associations between variables were described as odds ratios (OR) with their corresponding95% confidence intervals (CI). Correlations between continuous variables were assessed using the correlation coefficient. All tests were two-sided, and a p-value <0.05 was considered statistically significant. Analyses were performed using commercially available statistical software (GraphPad Prism, version 9.03 for Windows, San Diego, California, USA).

## Results

### Participants

A total of 168 patients (53.0% men) with a median age of 54 years [IQR 39.5–64.8] were enrolled. At the one-month follow-up, 151 (89.9%) participants were successfully contacted, and 137 (81.5%) were reached for the final evaluation (Figure 1).

**Figure 1:**
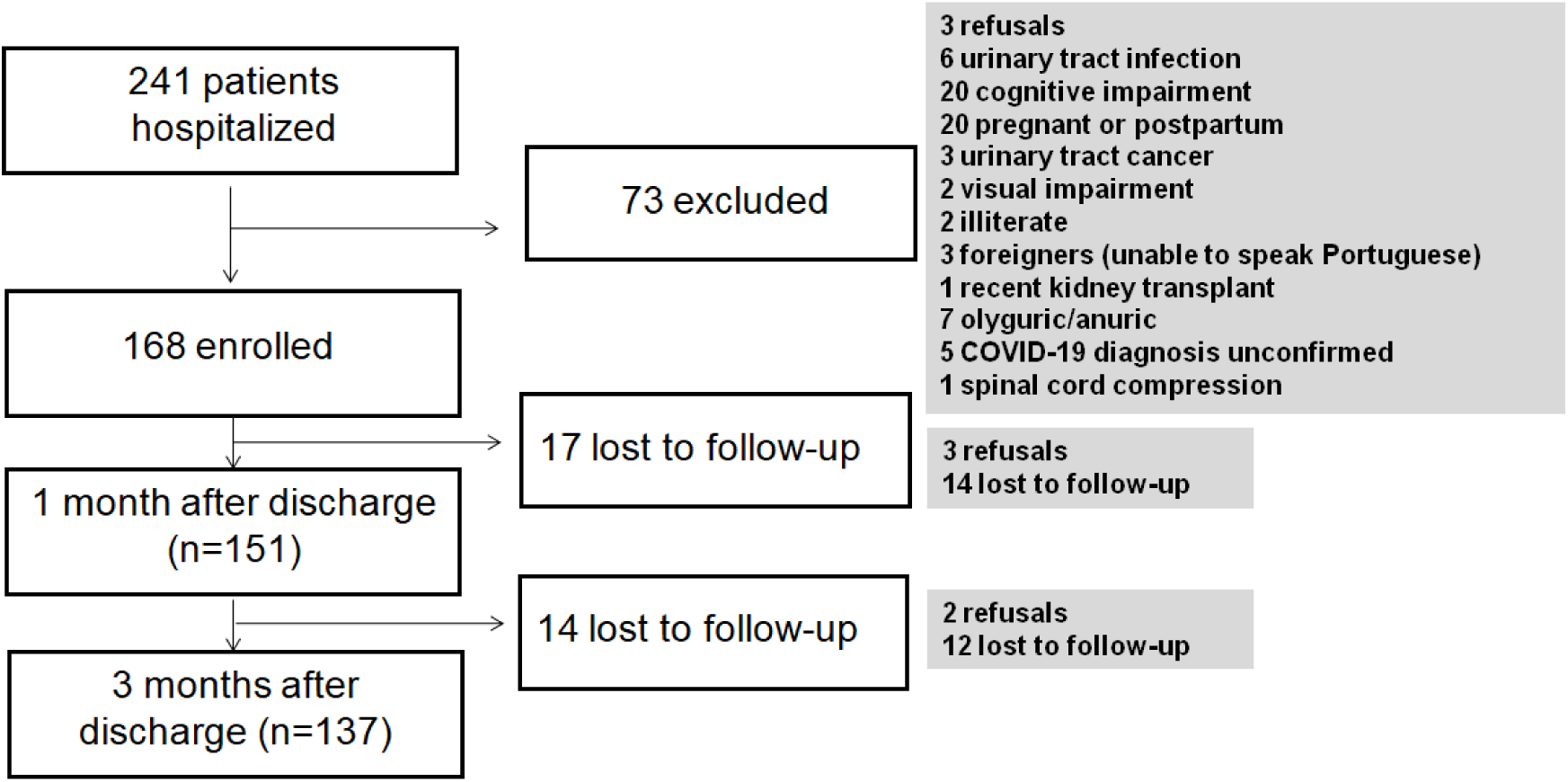
Study flow chart.

### Preexisting comorbidities, parameters of Covid-19 severity and vaccination status

The most prevalent comorbidities were systemic arterial hypertension (36.3%), obesity (33.9%), and diabetes mellitus (19.1%). Kidney transplant recipients constituted 17.3% of the cohort. Only 2 patients (1.2%) had an indwelling urethral catheter at the time of enrollment.

Regarding vaccination status, 96 participants (57.1%) were fully vaccinated, with a median of 85.5 days [IQR 41.8–149] between their last vaccine dose and symptom onset. Vaccines administered included ChAdOx1-S/nCoV-19 (39.6%), BBIBP-CorV (47.9%), BNT162b2 (11.5%), and JNJ-78436735 (1.0%). Men and women were comparable regarding preexisting comorbidities, COVID-19 severity, and vaccination status. Clinical data are summarized in Table 1.

**Table 1:** Demographic characteristics, preexisting conditions, and COVID-19 severity in the study population.

|  | **Women**<br>**(n=79)** | **Men (n=89)** | **Total (n=168)** |
| --- | --- | --- | --- |
| **Age (median)** | 55 [43.0-67.0] | 55.0 [41.0-64.0] | 54.0 [39.5-64.8] |
| **Pre-existing conditions** |  |  |  |
| Obesity | 28 (35.4%) | 29 (32.6%) | 57 (33.93%) |
| Arterial hypertension | 32 (40.5%) | 29 (32.6%) | 61 (36.31%) |
| Diabetes | 17 (21.5%) | 15 (16.9%) | 32 (19.05%) |
| Kidney transplant | 11 (13.9%) | 18 (20.2%) | 29 (17.26%) |
| Fully vaccinated | 48 (60.8%) | 48 (53.9%) | 96 (57.14%) |
| **Micturition** |  |  |  |
| Spontaneous | 78 (98.7%) | 88 (98.9%) | 166 (98.8%) |
| Bladder catheter | 1 (1.3%) | 1 (1.1%) | 2 (1.2%) |
| **COVID-19 severity** |  |  |  |
| Intensive care | 20 (25.3%) | 20 (22.5%) | 40 (23.81%) |
| Orotracheal intubation | 2 (2.5%) | 4 (4.5%) | 6 (3.60%) |
| Dyalisis | 0 (0%) | 01 (1.1%) | 1 (0.6%) |
| Pulm. Involv. $\geq 50\%^*$ | 7 (8.9%) | 12 (13.5%) | 19 (11.31%) |

\* Pulm. involv.> 50%: Pulmonary involvement on chest CT scans
Note: No statistically significant differences were observed between men and women for any of the variables presented

### Prevalence of LUTS and impact on quality of life

IPSS Scores: The initial median IPSS was 4.0 [IQR 2.0–8.3], with no significant difference between men and women. Moderate to severe LUTS were reported by 31.0% of participants, with storage symptoms being the most prevalent. Nocturia was reported by 53.6%, increased urinary frequency by 36.9%, incomplete emptying by 16.7%, slow urinary stream by 16.7%, intermittency by 11.9%, and straining by 8.9%.

ICIQ-OAB Scores: Urgency was reported by 51.9% of women and 33.7% of men (*p*=0.008) while UUI was reported by 36.7% of women and 15.7% of men (*p*=0.006). Overall, 36.7% of participants met the criteria for OAB, including 48.1% of women and 27.0% of men (*p*= 0.005).

ICIQ-UI SF Scores: Urinary incontinence was reported by 46.8% of women and 20.2% of men (*p*<0.001). SUI affected 38.0% of women and 3.4% of men (*p*<0.001). Post-micturition dribbling was reported by 12 participants (7.1%), with no significant difference between genders (*p*=0.522).

LUTS-related QoL impairment was reported by 13.9% of women and 14.6% of men (*p*=0.807). Among individuals with moderate-to-severe LUTS, 46.2% experienced a negative impact on QoL.

Pre-existing LUTS: Pre-existing LUTS were reported by 21.4% of participants, with no significant gender differences. Among these individuals, 69.4% felt their symptoms remained unchanged during the acute illness (58.8% of women and 78.9% of men), while 19.4% reported worsening symptoms (23.5% of women and 15.8% of men). Conversely, 11.1% experienced symptom improvement (17.6% of women and 5.3% of men). Among those with moderate to severe LUTS, 57.7% reported pre-existing symptoms, of whom 23.3% experienced symptom worsening during the acute illness.

### Follow-up

over the three months following hospital discharge, LUTS showed a general trend of improvement (Table 2).

**Table 2:**
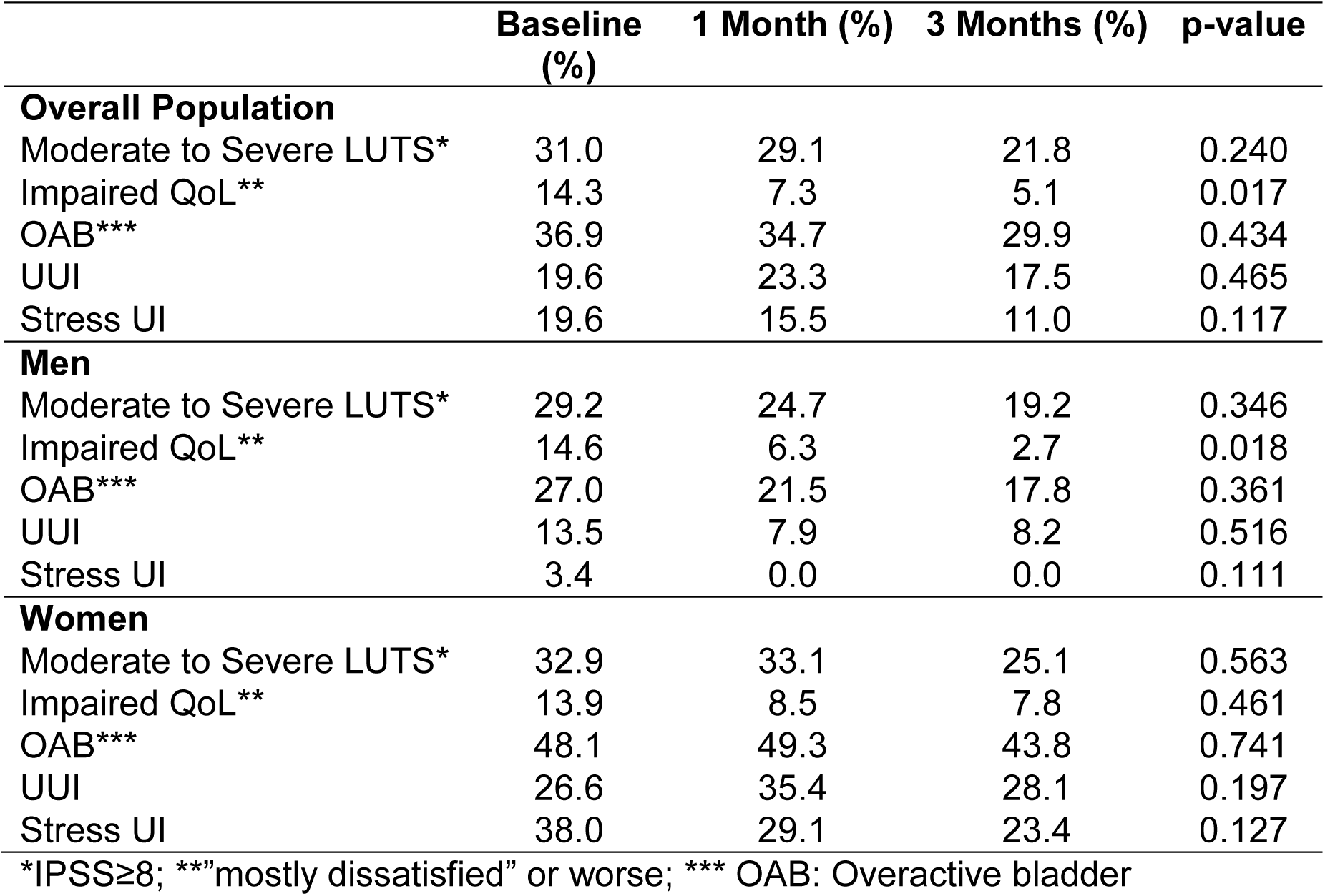
Prevalence and evolution of LUTS over time.

IPSS: The prevalence of moderate to severe LUTS decreased from 31.0% at baseline to 29.1% at one month and 21.9% at three months (*p*=0.240).

ICIQ-OAB: small decreases in the prevalence of OAB (36.9%at baseline vs 29.9% at three months; *p*=0.434) and UUI (19.6% at baseline vs 17.5% at three months; *p*=0.465) were observed over time.

ICIQ-UI SF: The prevalence of stress urinary incontinence (SUI) declined from 19.6% at baseline to 11.0% at three months (*p*=0.117).

LUTS-related QoL improved significantly over time.

### Association of Comorbidities, COVID-19 Severity and LUTS

Pre-existing conditions such as hypertension, diabetes, obesity, or kidney transplant status were not associated with LUTS during the acute phase of COVID-19. Similarly, parameters of COVID-19 severity, including ICU admission, orotracheal intubation, and lung involvement on CT scans, showed no correlation with the prevalence of LUTS.

### Impact of Vaccination on LUTS

Fully vaccinated individuals had an almost seven-fold risk of presenting with moderate to severe LUTS at baseline (OR 6.77, 95% CI 2.90–14.64; *p*<0.001), and a higher risk was still observed at three months post-discharge (OR 2.44, 95% CI 1.03–5.80; *p*=0.043).

When stratified by vaccine type, patients who received BBIBP-CorV (an inactivated-virus vaccine) were 2.23 times more likely to develop moderate to severe LUTS compared to those who received mRNA vaccines. The adjusted OR for BBIBP-CorV was 10.56 (95% CI: 4.13–26.9), while for mRNA vaccines it was 4.72 (95% CI: 1.84–12.03; *p*<0.001).

There was no difference in LUTS improvement over time between vaccinated and non-vaccinated individuals (Figure 3).

**Figure 2:**
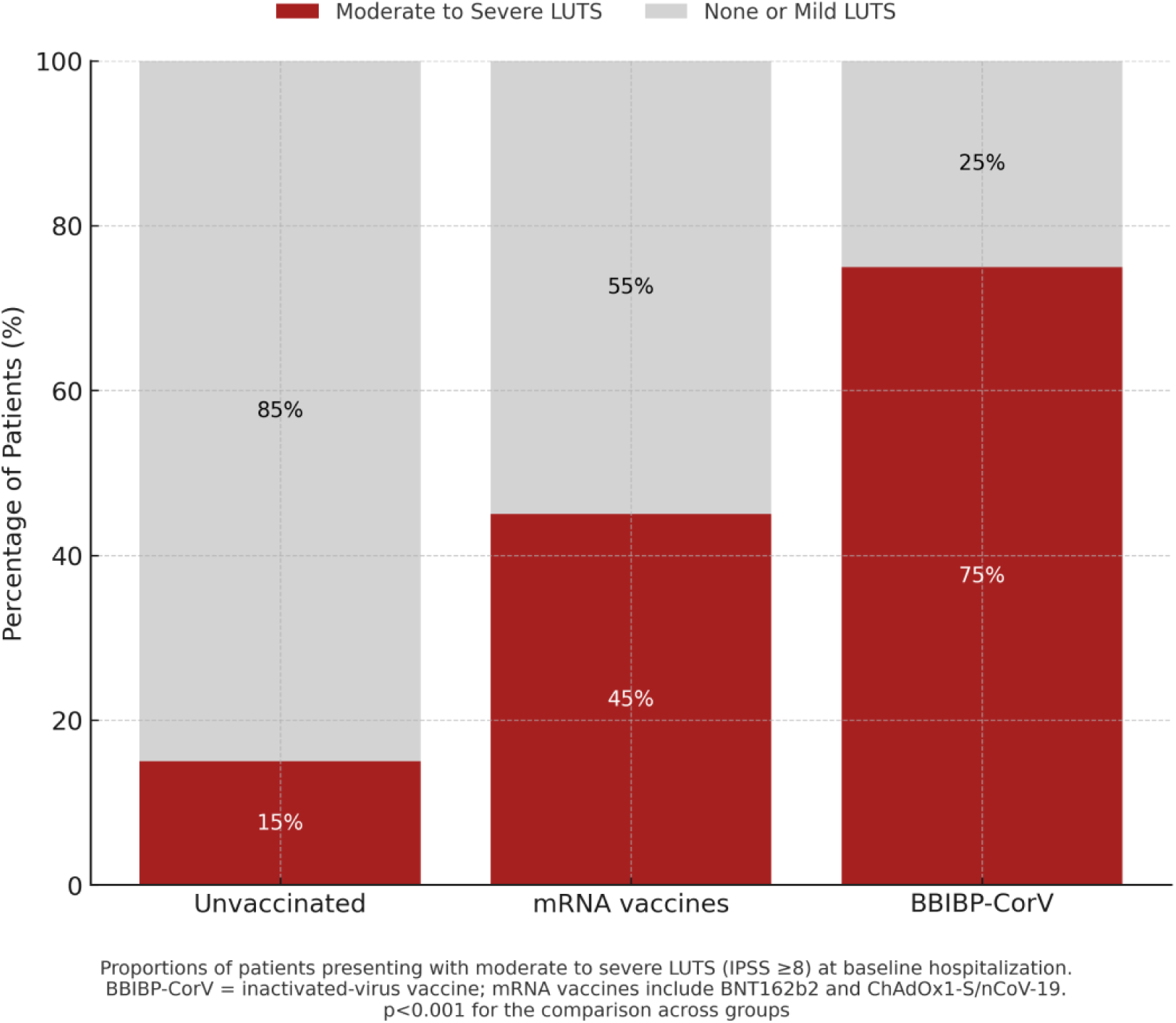
Proportion of Patients with Moderate to Severe LUTS by COVID-19 Vaccination Status and Vaccine Type.

**Figure 3:**
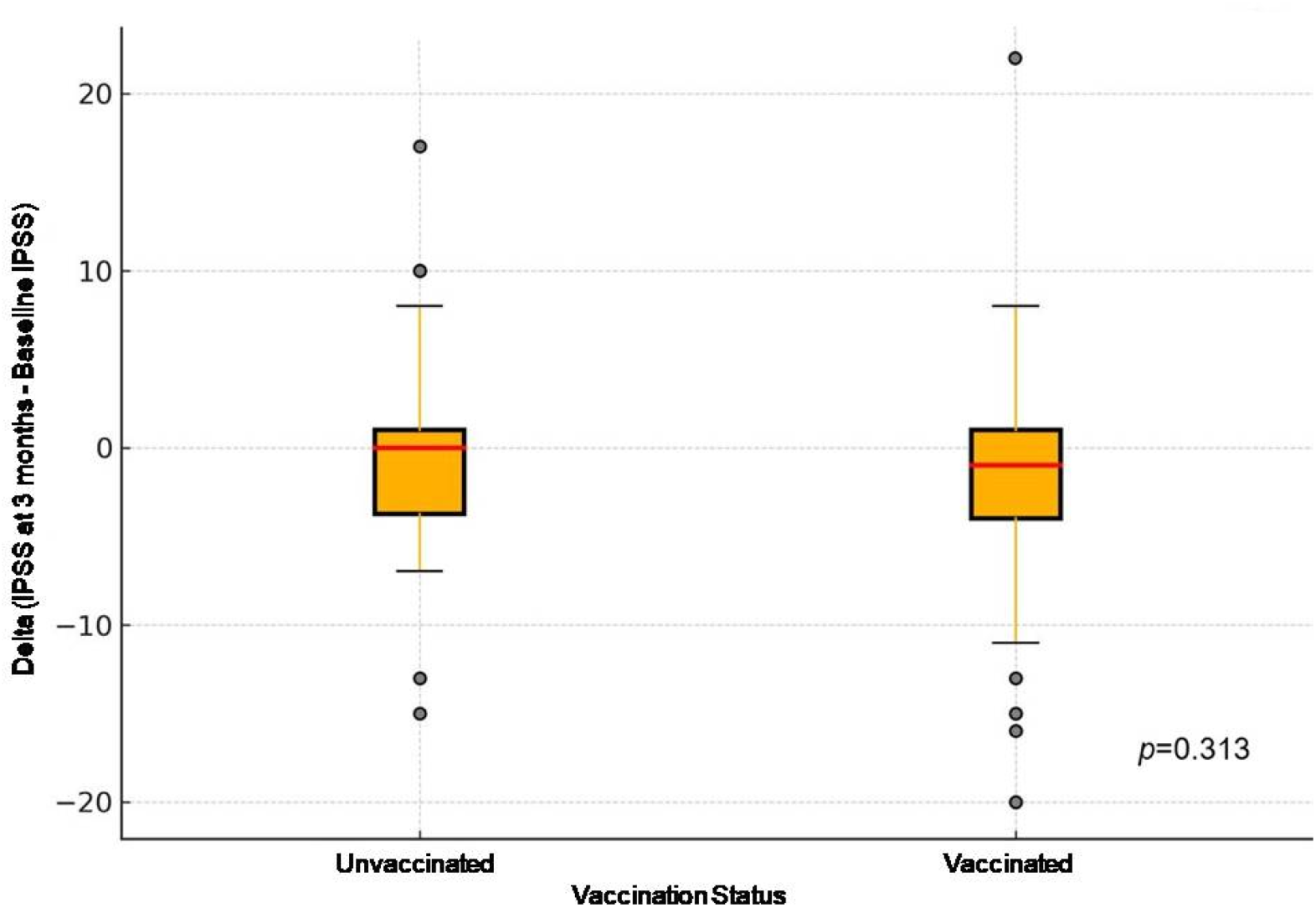
IPSS Variation at 3 Months According to Vaccination Status.

## Discussion

This study is the first to prospectively examine the prevalence, progression, and risk factors associated with LUTS in hospitalized COVID-19 patients, with follow-up extending to three months post-discharge. Key findings include: (1) a high prevalence of LUTS during acute infection, particularly storage symptoms; (2) LUTS improvement over time; and (3) a strong association between vaccination - especially inactivated-virus vaccines - and moderate to severe LUTS.

Women were more affected by storage symptoms, particularly urgency and incontinence. Similarly, a cross-sectional study involving 709 ambulatory COVID-19 patients reported higher rates of urgency, UUI, SUI, and frequency in women, whereas men more commonly reported straining [15]. However, that study lacked follow-up data.

We observed an overall improvement in LUTS over time. Although most comparisons did not reach statistical significance - likely due to the relatively small sample size - the 50% reduction in moderate to severe cases represents a clinically meaningful improvement, accompanied by a significant enhancement in LUTS-related QoL. Urinary incontinence, particularly stress incontinence, was frequently reported during the initial evaluation but gradually decreased by three months, likely reflecting the resolution of COVID-19-related coughing and sneezing.

In most cases, LUTS were either new-onset or worsened preexisting symptoms, suggesting both direct and indirect effects of COVID-19 on bladder function. The onset of de novo LUTS may reflect COVID-19-related bladder dysfunction mediated by inflammation, autonomic deregulation, or ACE2 receptor activity [16–18]. Conversely, exacerbation of preexisting LUTS may stem from physiological stress or secondary effects of hospitalization, such as altered mobility or medication use. Other studies reported higher LUTS rates in hospitalized COVID-19 patients [19, 20]. These findings emphasize the multifactorial nature of LUTS during and after COVID-19.

The observed improvement in LUTS over time contrasts with findings from our previous study on Long COVID, which reported a 42.4% prevalence of moderate to severe LUTS at six months post-COVID-19 [8].

Several factors may account for differences between the two studies. The earlier study was conducted with patients affected by a different virus variant before vaccines became available. Differing assessment tools were used to evaluate LUTS and may have influenced symptom reporting and prevalence rates. Also, the impact of Long COVID on broader aspects of health, including mental and physical well-being, may contribute to sustained LUTS at six months. This includes factors such as persistent inflammation, autonomic dysregulation, or reduced mobility, all of which could exacerbate LUTS over time. These findings suggest a complex interplay of acute and long-term effects of COVID-19 on LUTS and underscore the need for ongoing monitoring to better understand symptom trajectories and risk factors.

We found no association between LUTS and preexisting comorbidities among patients hospitalized with acute COVID-19, contrasting with our previous findings at six months post-discharge, where diabetes and hypertension were associated with LUTS. This discrepancy may reflect the masking of chronic disease effects during acute systemic illness. Other factors, such as vaccination status, vaccine type, viral variants, and treatment-related changes in lifestyle or medications, may also influence LUTS trajectories.

Markers of COVID-19 severity, such as ICU admission, orotracheal intubation, or pulmonary involvement, were not correlated with LUTS prevalence. Similarly, another study involving 94 hospitalized men found no correlation between COVID-19 severity and LUTS, as assessed by the IPSS questionnaire and chest CT findings [21]. In contrast, a study with 606 hospitalized patients reported an association between COVID-19 severity and LUTS, with higher IPSS scores observed in patients with more severe disease [19]. The conflicting results among these studies may be explained by differences in methodology, patient populations, or assessment tools. Additionally, it remains unclear whether distinct SARS-CoV-2 variants or demographic characteristics could influence the relationship between COVID-19 severity and LUTS.

An unexpected finding in our study was the increased risk of LUTS in fully vaccinated individuals, who were more likely to experience moderate to severe symptoms during the initial assessment and three months post-discharge. This association was particularly strong among recipients of the BBIBP-CorV vaccine compared to those who received mRNA or viral vector vaccines. These findings should be interpreted with caution given the study’s observational design and potential confounders, including vaccination timing, pre-existing conditions, and individual immune responses.

Prior studies have reported LUTS following COVID-19 vaccination. One study found that 13.4% of individuals experienced worsening storage LUTS after the first vaccine dose, particularly among those with pre-existing overactive bladder, suggesting an immune-mediated mechanism [22]. Another study in men with benign prostatic hyperplasia reported increased urgency, frequency, dysuria, and hematuria post-vaccination, without changes in PSA or prostate volume, also attributing symptoms to immune response [23]. While these studies did not evaluate symptom resolution or vaccine platform-specific effects, they highlighted a possible immunological basis for post-vaccine LUTS.

Additional reports describe rare cases of LUTS, such as urgency, following mRNA vaccination [24, 25]. Data from the VAERS system indicate that adverse urologic events are uncommon [26], with most reported cases associated with mRNA-1273 (Moderna), which was not used in Brazil during our study.

Our study builds on this evidence by being the first to evaluate LUTS in the combined context of vaccination and acute COVID-19. We found a significantly higher risk of moderate to severe LUTS in fully vaccinated individuals, especially those who received BBIBP-CorV (an inactivated-virus vaccine), compared to mRNA or viral vector vaccine recipients. These findings suggest that vaccine platform, timing of administration, and post-infection immune dynamics may influence LUTS presentation. These findings support further research into mechanisms by which vaccines may impact bladder function, which could inform safer, personalized vaccine guidance.

Our study has several limitations. Our cohort size was relatively small and included only patients who required hospitalization, limiting the applicability of our findings to milder cases of COVID-19. The study was conducted at a single tertiary hospital, which may limit the generalizability of our findings to other healthcare settings or populations. Variations in viral variants, vaccination status (including the number of doses, timing, and type of vaccine), and regional healthcare practices could also influence the rates of LUTS. In addition, data on pre-existing LUTS were based on retrospective self-reporting, which may introduce recall bias.

Nonetheless, our study has several strengths. It is the first to prospectively assess LUTS from acute COVID-19 through short-to-mid-term recovery and the first to explore associations between LUTS and vaccination status. The prospective nature of the study allowed for longitudinal data collection, with few patients lost to follow-up, enhancing the reliability of the findings. The use of validated LUTS questionnaires ensures robust and standardized symptom assessment. We also investigated whether LUTS were newly onset or had worsened after SARS-CoV-2infection. These features provide a unique and detailed perspective on LUTS in the context of COVID-19, offering critical insights into their prevalence, progression, and contributing factors.

In conclusion, LUTS were highly prevalent among patients hospitalized with COVID-19, particularly storage symptoms, and significantly affected quality of life. While symptoms improved over time, moderate to severe LUTS persisted in a notable subset. Our findings suggest a potential link between COVID-19 vaccination and LUTS severity, particularly with inactivated-virus platforms. Further prospective studies are needed to confirm these associations and guide management strategies.

## Data Availability

All data produced in the present study are available upon reasonable request to the authors.

